# Caregiver Mind-Mindedness Training as an Early Intervention for Social Anxiety in Children: A protocol for an RCT intervention

**DOI:** 10.1101/2024.11.23.24317842

**Authors:** Hiva Javadian, Mary E. Stewart, Minu Mathews, Andrew James Williams, Daniel Hale

## Abstract

**Background:** This paper describes a randomised controlled trial (RCT) protocol aimed at investigating the efficacy of caregivers’ Mind-Mindedness training as an early intervention for preschoolers with social anxiety. Mind-mindedness, a caregiver’s ability to recognise and respond to a child as an individual with their own thoughts, feelings, and intentions, is associated with secure attachment and socioemotional skills. While previous studies indicate brief Mind-Mindedness training may increase caregiver mentalisation in high-risk groups, rigorous research on the impacts of social anxiety in children is limited. Building on the well-established link between caregiver’s Mind-Mindedness and positive socioemotional outcomes in children, this study aims to bridge the existing research gap by directly testing the impact of Mind-Mindedness training on social anxiety.

**Methods:** This randomized controlled trial aims to recruit 100 caregivers of preschool-aged 4 to 7-year-old children with social anxiety from the UK and Iran. The caregivers will be randomly assigned to either a mind-mindedness training group (n=50) or a peer support (control) group(n=50). The mind-mindedness training will involve three online sessions focused on teaching strategies for using mind-minded comments, the program will consist of three online sessions, each lasting one hour and conducted over three consecutive weeks. The peer support (control) group will have access to a private online peer-support platform for sharing experiences. Measures of mind-mindedness, child social anxiety, attachment, and theory of mind will be assessed at baseline, post-intervention, and 3-month follow-up using established assessment tools.

**Discussion:** The study aims to evaluate the effectiveness of a mind-mindedness parental intervention for social anxiety in children and to uncover the potential mediating roles of attachment and theory of mind in the relationship between mind-mindedness and child anxiety. The cross-cultural design, involving participants from the UK and Iran, will offer valuable information on the cultural aspect of the intervention. The training group is hypothesized to lead to increased mind-mindedness and reduced child social anxiety versus a peer support (control) group. This research can establish evidence for mind-mindedness training as an early intervention approach for childhood social anxiety.

**Trial registration:** Prospectively registered on ClinicalTrials.gov (ID: NCT06657014; registered on 23rd October 2024) and on the Iranian Registry of Clinical Trials (IRCT) Version1(ID: 80088; approved on 2nd November 2024). These registrations promote transparency and ensure compliance with international standards for reporting clinical trials.

## 1. Introduction

### 1.1 Background and Rationale

Social anxiety impacts a significant number of children, particularly preschoolers, and has long-term mental health implications if left unaddressed (Costello et al., 2003; Hitchcock et al., 2009; Newman et al., 1996). Anxiety symptoms in young people have been exacerbated by the COVID-19 pandemic (Racine et al., 2021). Social anxiety is characterised by feelings of distress and avoidance of social settings due to a fear of negative evaluation (Beidel, 1991) which can impair the development of social relationships and pose challenges in adapting to school life (Strauss et al., 1987; Strauss et al., 1988). Anxiety levels often begin to present between the ages of 2-4, likely due to significant life changes such as starting preschool (Wang & Zhao, 2015; Zhou & Li, 2022). The preschool period is a critical stage in a child’s life, as it involves significant changes in the teacher, peers, and school environment, which can be challenging to navigate (Ladd, 2003; Pianta, 2007). Parenting style and attachment are strongly linked to childhood anxiety (Bogels & Brechman-Toussaint, 2006; Bowlby, 1969). Early intervention for preschoolers with social anxiety, through targeted approaches that focus on parenting and attachment, can help prevent symptoms from escalating into more severe disorders as the child develops (Albano, 2003; Bell-Dolan et al., 1990). The preschool years are therefore of critical importance for anxiety prevention efforts. While the impact of parenting style and attachment on a child’s anxiety is well-documented, interventions often focus exclusively on the child, neglecting the crucial role of caregiver behaviour (Bogels & Brechman-Toussaint, 2006).

One particular aspect of caregiver behaviour is Mind-Mindedness (MM). MM involves a caregiver’s ability to recognise and respond to a child as an individual with their own thoughts, feelings, and intentions (Fonagy & Luyten, 2009; Meins, 1997). Higher levels of MM are associated with secure attachment and positive socioemotional outcomes (Aldrich et al., 2021; Bernier, 2017; Meins, 1997; Sharp et al., 2006). There is little work focusing on whether caregiver’s MM relates to social anxiety in children or whether interventions which support MM in caregivers impact children’s social anxiety. However, there is some indirect evidence which may suggest that caregiver’s MM is associated with children’s social anxiety through attachment and Theory of Mind. For instance, caregiver MM is associated with attachment (Fonagy & Luyten, 2009; Meins, 1997) and ToM (Aldrich et al., 2021; Bernier & Dozier, 2003; Bernier, 2017; Foley et al., 2023).

Meins presents a link between lower MM and a child’s insecure attachment (Meins, 1998, 2011; Meins, 2001). Several interventions have been developed to enhance maternal MM, including video feedback and smartphone apps, all of which have demonstrated promise across diverse caregiver populations (Colonnesi et al., 2012; Larkin et al., 2019; Schacht et al., 2017). Importantly, it appears that caregivers’ socioeconomic status and child characteristics do not significantly impact MM levels, indicating that these interventions can be broadly applicable regardless of background or individual child traits (McMahon & Bernier, 2017; Meins, 2011). A relationship has been shown between maternal and paternal MM with child attachment security and theory of mind abilities (Bernier & Dozier, 2003; Foley et al., 2023).

Based on attachment theory, early experiences with caregivers shape internal working models that affect future relationships (Bowlby, 1969). Thus, children who are securely attached are better able to understand different perspectives within relationships (Berlin et al., 1995). Insecure attachment is a risk factor for internalising problems such as anxiety (Brumariu & Kerns, 2010; Colonnesi et al., 2011) while secure attachment is associated with appropriate mind-related comments and fewer non-attuned comments (Arnott, 2007; Meins, 2001). Previous studies have indicated that parent cognitions may play a vital role in mediating the relationship between parent and child difficulties (Dix et al., 1986; Sadler et al., 2006; Sharp et al., 2006; Wheatcroft & Creswell, 2007). Some research suggests that MM might be a better predictor than the sensitivity of attachment status (E Meins, 2002) at least be a prerequisite for sensitivity (Laranjo et al., 2008). Therefore, focusing on MM in interventions may improve outcomes for families.

Maternal MM has been found to be predictive of children’s Theory of mind (ToM), the ability to understand and attribute mental states to oneself and others (Bernier et al., 2010; Meins, 1999; Wellman, 1992). Mothers who describe their children in more mental terms have children who perform better on tasks that require an understanding of false beliefs (Hughes et al., 2018; Meins, 1998). Moreover, higher ToM abilities are linked to secure attachment in preschoolers (Laranjo et al., 2014; Meins et al., 2018). On the other hand, poor ToM abilities are associated with childhood anxiety disorders and negative interpretive biases (Hezel, 2014; Moldovan & Visu-Petra, 2022).

This study will focus on examining whether training caregivers in MM can reduce social anxiety in preschoolers by improving attachment security and ToM abilities. Understanding these pathways from MM to childhood anxiety can help inform preventative interventions targeting early socio-cognitive development. Therefore, the proposed mechanisms in this study are attachment and theory of mind. MM strongly predicts child attachment security (Laranjo et al., 2008), which provides an anxiety buffer. Insecure attachment is linked to increased internalizing symptoms (Colonnesi et al., 2011). Caregiver’s MM also forecasts children’s theory of mind abilities (Meins, 2013).

Parental mentalization (MM) is strongly linked to secure attachment in children. Interventions that target MM could improve attachment security. Future research should prioritize MM as a key intervention target to address gaps in how attachment behaviours are passed from parent to child, especially when parents struggle to understand and respond to their child’s emotional needs. Consistent parental behaviours, such as emotional responsiveness, promote secure attachment in children. Interventions, such as training programs to enhance parents’ ability to perceive and interpret their child’s emotions, can help bridge these gaps, supporting children’s emotional and social development (Arnott, 2007; Atkinson & Goldberg, 2003; Bernier & Dozier, 2003; Tansy M. Walker et al., 2012).

Various interventions have been developed to enhance MM, ranging from video feedback to smartphone apps, demonstrating promise in diverse caregiver populations (Colonnesi et al., 2012; Larkin et al., 2019; Schacht et al., 2017). Upon closer examination of these interventions, it becomes evident that their effectiveness largely depends on addressing the unique needs of caregivers. Factors such as accessibility, ease of use, and the relevance of the content play crucial roles in determining the success of each intervention format. By enhancing caregivers’ MM, they are better equipped to cultivate secure attachments and establish a nurturing environment, and they can significantly decrease the symptoms of social anxiety in their children.

Various interventions have been conducted by researchers to explore the link between child development and caregivers’ mind-related comments. Schacht et al.(2017) reported that a single-session video-feedback intervention was effective in reducing non-attuned comments (Schacht et al., 2017). This intervention proved to be beneficial for mother-infant interactions even up to the second year of life. Another study conducted by Larkin et al. developed the BabyMind app, which was successful in increasing mothers’ MM even among vulnerable groups, such as adolescent mothers (Larkin et al., 2019). Walker’s (2012) research emphasized the importance of MM in preschool clinical interventions (T. M. Walker et al., 2012).

This paper proposes an RCT of MM training for caregivers of socially anxious preschoolers. This study emphasizes the need to adapt interventions for diverse populations. Research indicates culture shapes caregiving approaches, parenting behaviours, and attachment styles (Arnett & Arnett, 2015; Keller, 2013). Some studies reveal differences in MM across Western and Eastern cultures. For instance, British parents exhibit higher MM towards children compared to Chinese parents, but both show positive links between MM and child theory of mind (Hughes et al., 2017). Research indicates that Western mothers engage in more mental state talk with children than Eastern mothers (Doan & Wang, 2010). Collectivist values in Eastern cultures prioritizing hierarchy and conformity versus individuality in the West impact manifestations of MM in parent-child interactions (Kağıtçıbaşı, 2007; Keller et al., 2007). According to some research traditional gender norms can also affect father-daughter relationships and emotional attunement in Persian families specifically (Golian Tehrani et al., 2015; Mousavi et al., 2020). For instance, in a study comparing parenting practices in the UK and India, they found notable differences: UK mothers made more mind-minded comments, while Indian mothers issued more instructions and positive comments, reflecting cultural priorities (Bozicevic et al., 2023). These differences highlight the influence of cultural contexts on parenting practices. Recognizing and understanding these variations can guide the development of interventions sensitive to cultural contexts, ultimately promoting healthier outcomes for children worldwide(Arnett & Arnett, 2015; Bornstein, 2006).

## 2. Materials and Methods

### 2.1 Objectives

The main aim of this study is to evaluate the efficacy of the caregiver’s MM training in reducing social anxiety symptoms among preschool children. Additionally, to examine how culture (Iranian and British) moderates the relationship between caregiver’s MM training and reductions in social anxiety symptoms among preschool children.

### 2.2 Trial design

This study protocol outlines the design and methodology of a randomized controlled trial aimed at investigating whether MM training for caregivers can effectively reduce social anxiety symptoms in preschoolers by employing a cross-cultural approach involving caregivers from the UK and Iran. Participants will be randomly assigned to either the MM training intervention group or a peer support (control) group. Randomization will be carried out separately for each country’s sample. Stratified random sampling will ensure a balanced allocation within each country’s participants.

### 2.3 Study setting

The study will be conducted in two countries: the United Kingdom and Iran. The intervention and all measurements will be held online.

### 2.4 Participants

The study aims to recruit 100 primary caregivers (aged 18-60 years) of preschool-aged children (aged 4-7 years) who exhibit symptoms of social anxiety. Eligible participants must reside in either the United Kingdom or Iran. Caregivers whose children have existing clinical diagnoses or are currently receiving other forms of treatment or support will not be included in the study.

Recruitment strategies will include advertisements in preschools and nurseries, word-of-mouth referrals, and recommendations from teachers. Participants will be selected based on observable social anxiety symptoms, with the goal of creating a homogeneous sample. The sample will be balanced, ensuring an equal distribution between the two countries. Due to the nature of the intervention, participant blinding is not possible, as caregivers will need to actively engage in either mindfulness meditation training or peer support groups and will be aware of their group assignments. However, the assessors responsible for data collection and analysis—including those evaluating mindfulness meditation, child social anxiety, attachment, and theory of mind—will remain blinded to the group allocations to ensure an unbiased evaluation of outcomes.

To enhance participant retention and reduce attrition, several strategies will be implemented. These will include clear communication protocols, flexible scheduling options, and modest incentives for completing assessments. Participants will receive systematic reminders about their sessions, and a dedicated support structure will be available throughout the duration of the study. Additionally, data from participants who deviate from the protocol will be included in the intention-to-treat analysis. This approach will help preserve the integrity of the randomization process while minimizing bias and ensuring that the findings are practically applicable and generalizable.

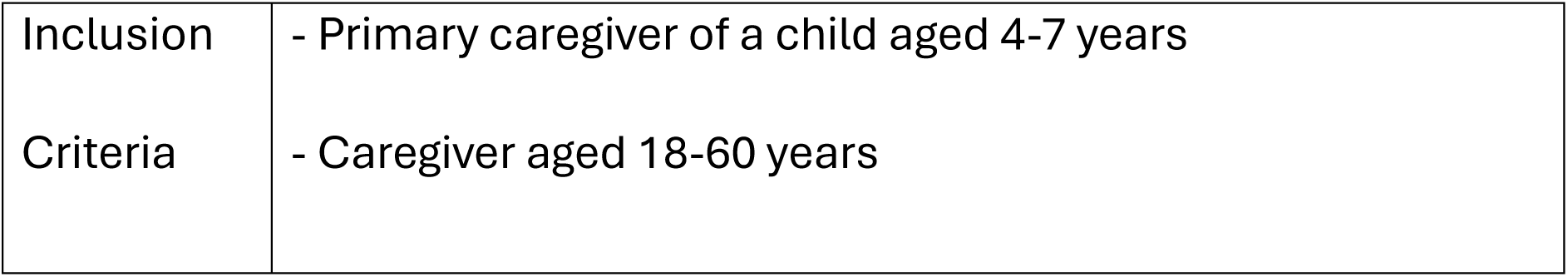
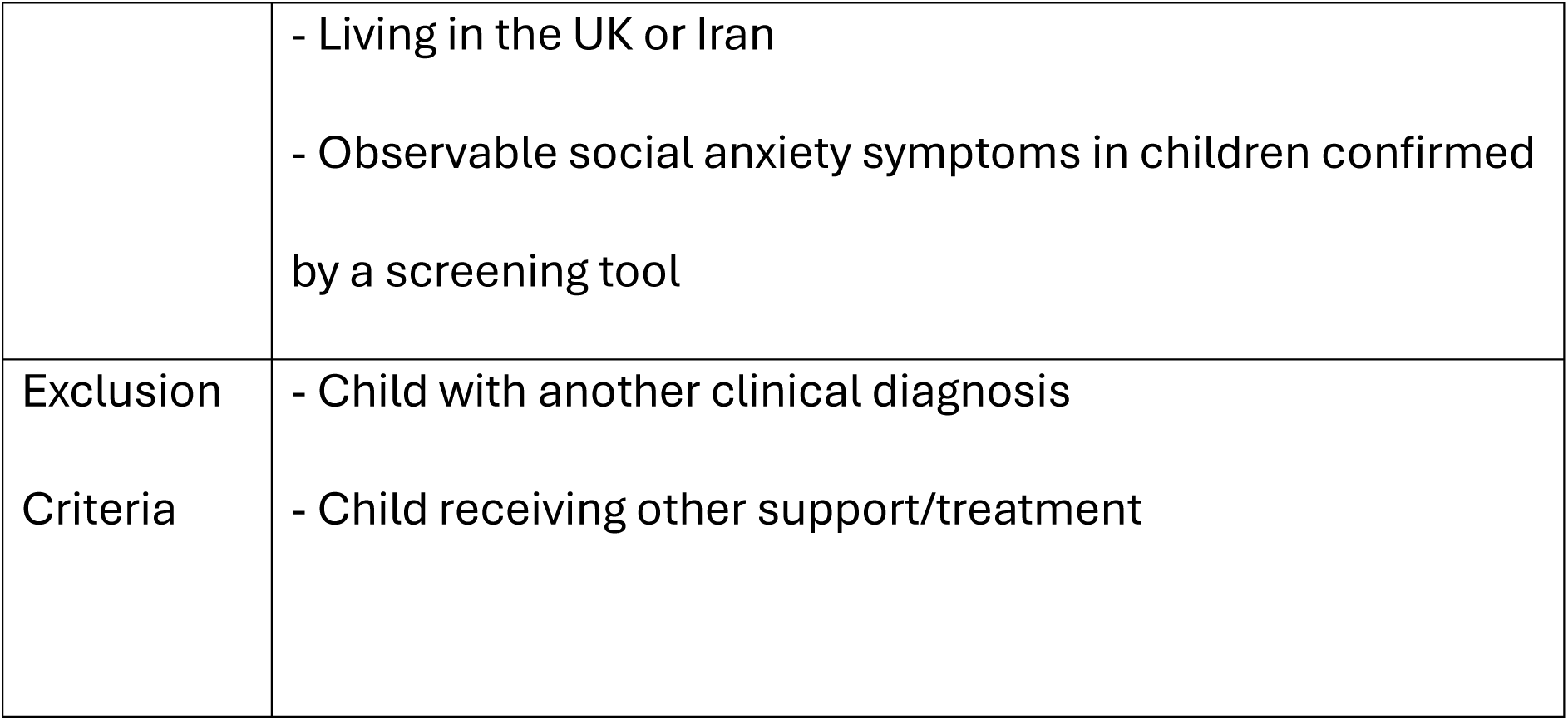

### 2.5 Procedure

Participant allocation will be carried out using a computer-generated randomization sequence through Random Allocation Software 2.0, which will assign caregivers to either the mind-mindedness (MM) training intervention group (n=50) or the peer support control group (n=50). The randomization will be stratified by country (UK and Iran) to maintain a balanced allocation within each cultural context.

Data collection will take place at three time points: baseline (T0), post-intervention (T1), and 3-month follow-up (T2). This will utilize validated assessment measures, including the MM Coding Manual 2.2(Meins, 2015), for mind-mindedness, the Spence Children’s Anxiety Scale(Spence, 1997), for assessing child social anxiety, the Attachment Insecurity Screening Inventory (Spruit et al., 2018), for attachment assessment, and the Theory of Mind Task Battery(Hutchins et al., 2012) for evaluating the theory of mind.

Additionally, demographic information (such as age, gender, socio-economic status, and cultural background) will be collected to describe the study population and control for potential confounding variables. Program adherence will be monitored through attendance records and weekly caregiver reflections on their experiences.

The informed consent process will follow institutional guidelines, ensuring participants receive detailed study information in their preferred language (English or Farsi). Participants will have opportunities to ask questions and will receive a clear explanation of their right to withdraw from the study. The Principal Investigator (PI) is responsible for obtaining informed consent from all potential trial participants.

Comprehensive data protection measures will be implemented, including secure storage of consent forms and assessment data in encrypted, password-protected files. Unique participant identifiers will be assigned, and access will be restricted to authorized research personnel only. All data management procedures will comply with GDPR and local data protection regulations. The final dataset will be accessible only to the research team and will be stored according to Heriot-Watt University’s data retention policy. This thorough approach to randomization, data collection, and protection ensures both scientific validity and participant confidentiality throughout the study.

**Fig1.**
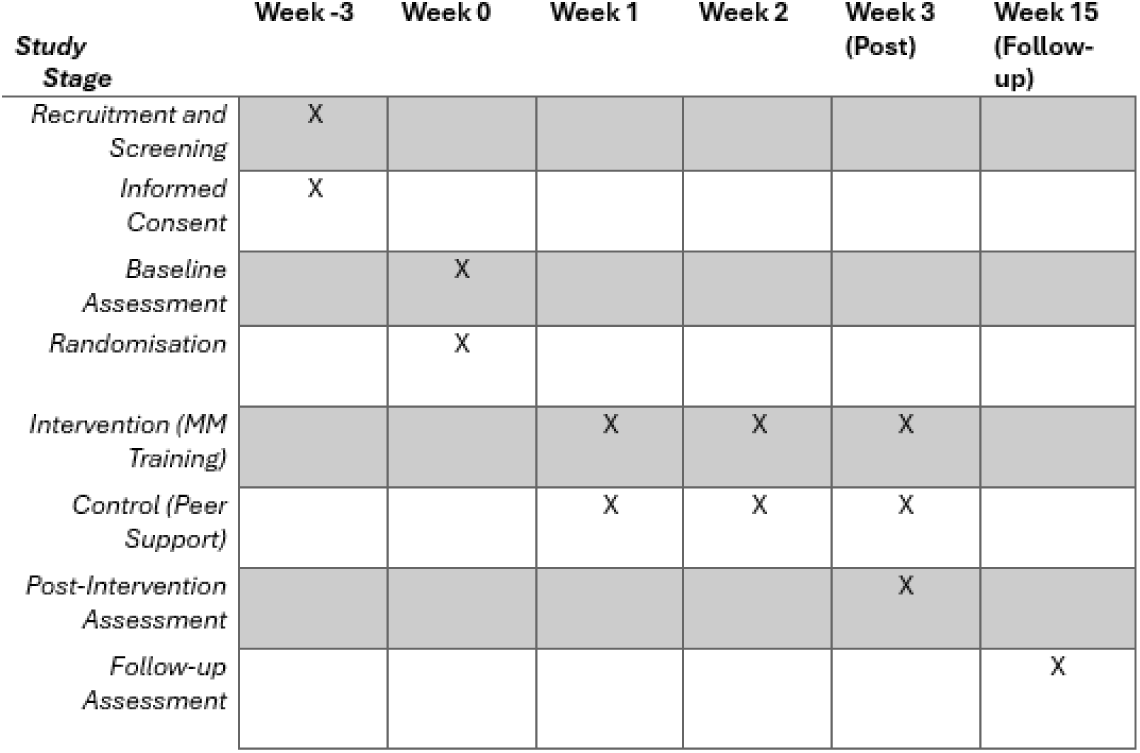
Participants’ Timeline.

### 2.6 Intervention

This study aims to reduce social anxiety in preschoolers by tailoring an MM intervention that targets parents to alleviate social anxiety levels in their children. Collaborating with parents and caregivers will result in a more effective and tailored solution that resonates with their practical needs and experiences. Their input can lead to a more feasible and impactful study design, refining eligibility criteria, recruitment, and data collection procedures. A participatory co-design was conducted in order to strengthen the quality and outcomes of this research. The intervention will be identical in both countries, with the only difference being the language used for the program. All other elements, such as the structure, content, delivery methods, and materials of the MM training program, will remain uniform across both locations.

The MM training program will consist of three online sessions, each lasting one hour and conducted over three consecutive weeks. The training sessions will be conducted through Zoom, and participants will receive a unique link to access the platform securely. This training is designed to equip caregivers with strategies for attuned, mentalizing caregiving, focusing on enhancing their ability to understand and respond to their child’s emotional and mental states.

The first session will introduce the concept of mind-mindedness and its importance in caregiving. It will include psychoeducation on the development of social anxiety in children and how caregiver responsiveness plays a crucial role. Caregivers will also be introduced to basic emotion coaching techniques, which will be explored in more depth in the second session. The second session will focus on teaching and practising these emotion coaching techniques, helping caregivers recognize, validate, and guide their children’s emotions in a supportive manner. The final session will deepen the caregivers’ understanding through reflective discussions and advanced role-play scenarios that address common challenges in caregiving with socially anxious children.

All three sessions will include video examples demonstrating effective and ineffective approaches, as well as role-play exercises where caregivers can practice these techniques with guided feedback. The role-plays will be led by trained child psychologists. They will give immediate feedback and answer questions to ensure that caregivers are effectively practicing mentalizing caregiving. Throughout the training, caregivers will receive support materials and reminders to track their progress and reflections. Participation will be tracked through attendance records and the completion of reflection activities to make sure that participants are following the program guidelines.

The peer support (control) group group will access a private online group for the exchange of experiences and coping strategies related to the child’s anxiety, without direct MM training.

The comprehensive set of measures includes assessments of MM, child anxiety scales, and evaluations of attachment and theory of mind.

Measures of MM will be coded from caregiver descriptions of the child using the MM Coding Manual 2.2 (Meins, 2015). Child anxiety will be assessed with the Spence Children’s Anxiety Scale (Spence, 1997).

Attachment will be measured using the Attachment Insecurity Screening Inventory (Spruit et al., 2018). Theory of mind will utilize the Theory of Mind Task Battery (Hutchins et al., 2008). The three-day MM training program incorporates psychoeducation, role-play exercises, and practical sessions, while the peer support (control) group serves as an active control condition.

**Fig2.**
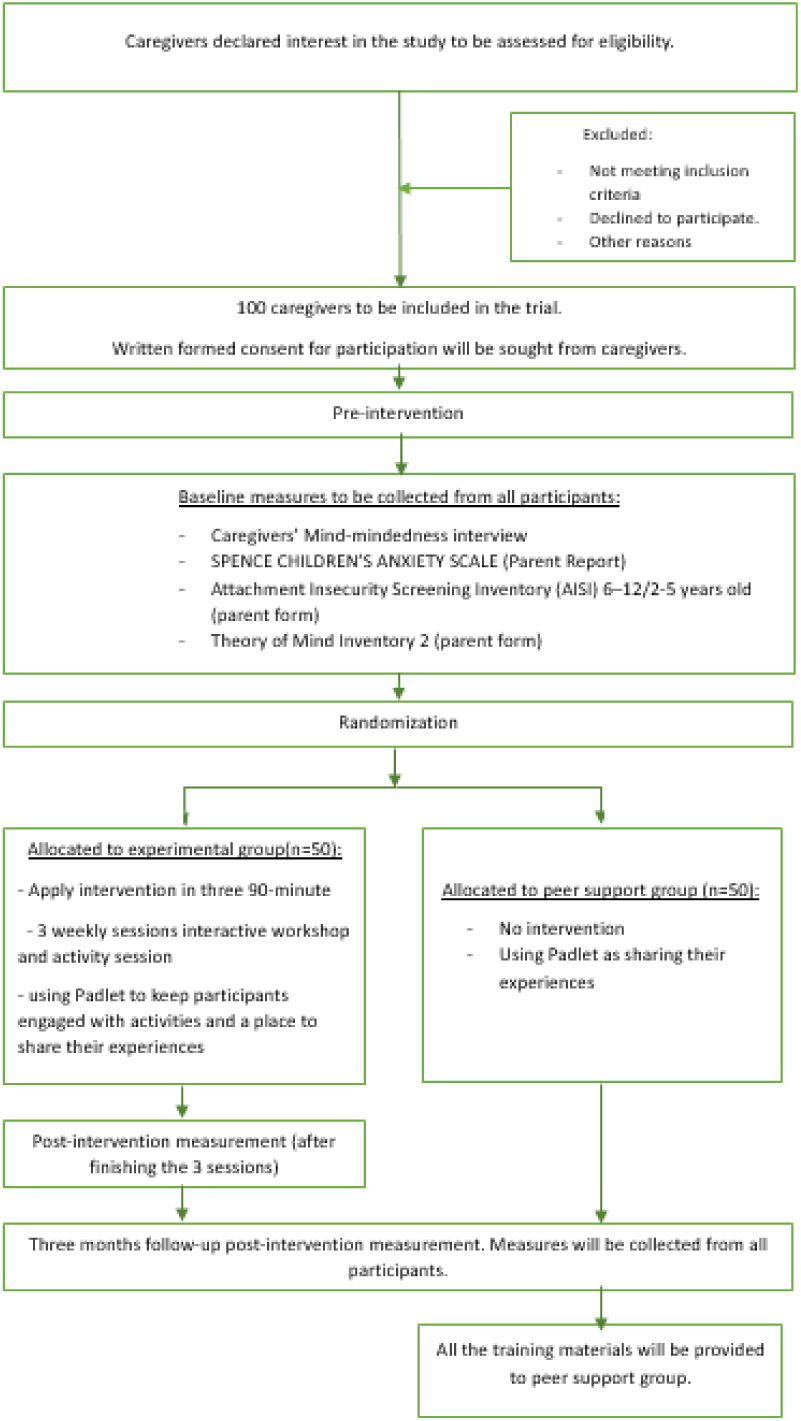
RCT design.

### 2.7 Statistical methods

#### 2.7.1 Sample Size and Power

The study aims to recruit 100 primary caregivers, with 50 from the UK and 50 from Iran. This sample size was determined through power analysis using G*Power 3.1, which indicates that it provides 80% power to detect medium-to-large effects (Cohen’s d ≥ 0.5) at an alpha level of 0.05 for the primary outcome of child social anxiety in repeated measures ANOVA. We have accounted for an anticipated attrition rate of 20%. The balanced allocation of participants between the two countries allows for robust cross-cultural comparisons while maintaining adequate power for within-group analyses.

#### 2.7.2 Primary Analysis

The primary analysis will adhere to intention-to-treat principles, employing a difference-in-differences (DiD) approach to evaluate the effects of the intervention. Missing data will be addressed using multiple imputations with 20 datasets. The main outcomes will be analyzed using a 2 (group: intervention vs. control) × 2 (culture: UK vs. Iran) × 3 (time: baseline, post-intervention, 3-month follow-up) repeated measures ANOVA, applying Greenhouse-Geisser corrections for any violations of sphericity where necessary. Effect sizes will be reported as partial η² with 95% confidence intervals.

#### 2.7.3 Secondary Analyses

Mediation Analysis: Path analysis using structural equation modelling will test whether attachment security and theory of mind mediate the relationship between MM training and child anxiety reduction. Model fit will be assessed using standard indices (CFI > 0.95, RMSEA < 0.06, SRMR < 0.08).

Cultural Moderation: Moderation effects will be tested using hierarchical linear modelling with culture as a level-2 predictor. Interaction terms will be decomposed using simple slope analysis.

Exploratory Analyses: Pearson correlations will examine relationships among key variables (MM, anxiety, attachment, ToM), with Bonferroni corrections for multiple comparisons (α = 0.05/number of comparisons).

#### 2.7.4 Data Management

Data will be collected electronically using REDCap, ensuring standardized collection and secure storage. Quality control includes double data entry for 20% of cases to verify accuracy,

automated range checks and validation rules, weekly backup procedures and regular data integrity audits. All analyses will be conducted using Stata 17.0 (StataCorp, College Station, TX) with a significance level of p < 0.05 (two-tailed). Results will be reported following CONSORT guidelines for randomized trials.

#### 2.7.5 Safety Monitoring

Given the low-risk nature of the intervention, formal interim analyses and stopping rules are not planned. However, adverse events will be monitored continuously and reported according to standard protocols. Data security will be maintained through encryption, password protection, and restricted access to authorized personnel only.

## 3. Discussion

This study will contribute to the understanding of MM in parenting and its potential as a clinical intervention for preschoolers with social anxiety. The literature underscores the need for experimental trials to determine whether interventions in MM can effectively reduce social anxiety, given the current lack of such studies, while also acknowledging the associations between mentalization, attachment, and theory of mind.

The rationale behind this study lies in the existing literature that highlights the crucial role of MM in fostering positive socioemotional outcomes in children (Aldrich et al., 2021; Bernier, 2017). This study builds upon this foundation, intending to provide valuable insights into the effectiveness of an MM training program for caregivers of preschoolers with social anxiety. The envisioned randomized controlled trial design incorporates a comprehensive curriculum, including psychoeducation, role-play exercises, and practical sessions, with the aim of enhancing caregivers’ ability to recognize and appropriately respond to their child’s internal states. While the outcomes of the trial remain to be seen, the study’s design aligns with the intent of offering an innovative early intervention approach.

The cross-cultural component of the study is particularly noteworthy. The cross-cultural investigation enriches the study by exploring societal and cultural influences on MM, with implications for the development of culturally sensitive family-based treatments. Comparing an individualistic Western culture (UK) to a more collectivist Middle Eastern culture (Iran) will provide new insights into cultural variations in MM and impacts on intervention effectiveness. Exploring culture as a moderator can inform culturally sensitive applications (Hughes et al., 2018; Martinez & Mahoney, 2022; Mishu et al., 2023; Okoniewski et al., 2022; Rathod et al., 2018; Strand et al., 2019). The co-design process, involving caregivers in shaping the intervention, adds a participatory dimension that can enhance the relevance and impact of the study.

The research has potential implications for clinical practice and future research. If the trial demonstrates positive outcomes, it could pave the way for integrating MM training into existing parenting programs as a targeted approach to alleviate social anxiety symptoms in preschoolers.

Additionally, the study’s findings may prompt further exploration of the cultural variations in parental reflections on children’s internal states and their implications for intervention design. The findings are expected to inform clinical practice and contribute to the growing body of research on early interventions for childhood mental health. As previous studies (see (McMahon & Bernier, 2017) for a review) suggest links between MM and enhanced attachment security and theory of mind, the training may alleviate social anxiety by strengthening these protective socio-emotional factors. However, direct evidence establishing MM training as an anxiety intervention is limited. The proposed MM training program, if proven effective, could offer a valuable addition to the arsenal of strategies aimed at promoting positive mental health outcomes in preschoolers. The study’s design, cultural sensitivity, and participatory approach in shaping the intervention position it as a potential catalyst for advancing research and clinical practices in the realm of child mental health.

### 3.1 Trial status

Ethical approval for the study was granted by the Social Sciences Ethics Committee of Heriot-Watt university in September 2024. Recruitment will commence on 16th September 2024. This study is expected to be completed by February 2025.

## Data Availability

All relevant data are within the manuscript and its Supporting Information files. Receutment has not started yet.

## 4. Acknowledgement

I would like to convey my sincere gratitude to my primary supervisor, Daniel Hale, for his invaluable guidance and unwavering support throughout this project. I am also appreciative of the insightful comments and feedback from Mary E. Stewart and Minu Mathews. Additionally, I would like to extend special thanks to Andrew James Williams for his contributions and assistance.

## Appendix 1

### 1 Information Sheet

#### PARTICIPANT INFORMATION SHEET

PROJECT TITLE: Caregiver Mind-Mindedness training as an Early Intervention for Socially Anxious Children

#### INVITATION

You are being invited to take part in a research study for an intervention for children’s social anxiety. You must be the primary caregiver, responsible for the daily care of a child aged 4-7 with social anxiety symptoms, age 18-50. You must also have access to the internet. The research is supervised by Daniel Hale, Mary E. Stewart and Minu Mathews. The investigator is Hiva Javadian. The project has been approved by the School of Social Sciences Ethics Committee at Heriot-Watt University.

#### WHAT WILL HAPPEN

If you decide to participate, you will first have a brief interview lasting up to 30 minutes, which will be audio recorded, and 3 questionnaires to fill in. Following this, you will be randomly assigned to one of two groups:

##### 1- Mind-Mindedness (MM) Training Group

Participants in this group will attend a training program designed to enhance their understanding and responsiveness to their child’s emotional and mental states. The training includes psychoeducation, emotion coaching, video examples, reflective discussions, and role-play exercises. The aim is to promote attuned and mentalizing caregiving behaviours.

Commitment: Three 1-hour sessions held over three weeks.

##### 2- Peer Support Group

Participants in this group will have access to a private online platform. This space allows caregivers to share experiences and strategies related to managing their child’s anxiety, fostering a supportive community environment.

Commitment: Time spent engaging with the platform will vary based on personal participation.

Regardless of your group, you will need to undergo assessments before the intervention, immediately after the intervention, and three months after the intervention. These assessments, which will each take about an hour, are designed to measure mind-mindedness, your child’s anxiety levels, attachment, and theory of mind.

Note: If you are assigned to the peer support group, you will still receive the complete set of training materials used in the mind-mindedness training group after the three-month follow-up assessment. This ensures that all participants have access to the valuable content of the training, regardless of their initial group assignment.

#### AIM OF THE PROJECT

The aim of the study is to investigate the effectiveness of caregivers’ mind-mindedness training in reducing social anxiety symptoms among preschool-aged children. By assessing the impact of a targeted intervention on caregiver behavior and its effects on child social anxiety, this research aims to contribute to the development of early interventions for enhancing child well-being and promoting mental health outcomes.

Moreover, this study will explore the potential role of attachment and theory of mind as underlying mechanisms for the observed changes in social anxiety symptoms. This study will focus on how cultural differences between Iran and the UK may moderate this relationship.

What is Mind-Mindedness?

Mind-mindedness refers to the ability to recognize and consider a child’s thoughts, feelings, and intentions. In this study, Mind-Mindedness training aims to improve caregivers’ ability to understand and respond to their child’s inner experiences, which may help reduce the child’s social anxiety.

Please be aware that participants will be assigned randomly to either the Mind-Mindedness training or the peer support group to ensure a fair and unbiased evaluation of the interventions.

#### TIME COMMITMENT

Mind-Mindedness Training Group: Attend three 1-hour training sessions over three weeks.

Assessments: Complete assessments at three time points (baseline, post-intervention, three-month follow-up), each taking about one hour.

Peer Support Group: Engage with the online platform, with the time commitment varying based on your level of participation.

Your participation in this study is greatly appreciated. It could make a significant difference in understanding and addressing social anxiety in young children.

#### PARTICIPANTS’ RIGHTS

You may decide to stop being a part of the research study at any time without explanation. You have the right to ask that any data you have supplied to that point be withdrawn until the data is fully anonymised. Full anonymisation of your data will only happen once the data has been analysed. You have the right to omit or refuse to answer or respond to any question that is asked of you.

#### BENEFITS AND RISKS

There are no known benefits or risks for you in this study.

#### COST, REIMBURSEMENT AND COMPENSATION

Your participation in this study is voluntary; there is no compensation for your participation in this study

#### PRIVACY AND CONFIDENTIALITY

All data will be fully anonymized during the transcription process. Any identifying details, such as names, addresses, and places of work, will be removed or destroyed to ensure confidentiality.

Heriot-Watt University is the data controller for the personal data collected in this project. We will collect and use your personal data for this project only with your consent. You can withdraw your consent at any time until the data is fully anonymised by contacting the researcher, supervisor or the data protection team.

We will keep your personal data securely and ensure that no one will link the research data you provide to any identifying information you may supply. Once we have analysed the information you provide, we will completely anonymise your personal data so that it will not be possible to identify you from any information in the remaining dataset. After the project ends, we may use the anonymous dataset for research outputs such as articles and conference presentations.

If you would like to know more about what Heriot-Watt University does with your personal data and your rights under privacy law, please visit our data protection web pages at https://www.hw.ac.uk/uk/services/information-governance/protect/privacy-and-your-data-rights.htm or contact our Data Protection Officer by email at.

#### FOR FURTHER INFORMATION

Daniel Hale will be glad to answer your questions about this study and provide additional information on results if requested. You may contact them at.

### 2. Participant Consent Form

By signing below, you are agreeing that:

- You have read and understand the Participant Information Sheet and Consent Form
- You understand that there are no expected potential risks to you in your participation
- You are taking part in this research study voluntarily (without coercion or remuneration).
- You consent for any personal data1 collected to be used as part of this study
- Any questions you may have about your participation in this study have been answered satisfactorily

Personal data refers to information in any format about an individual who can be identified directly or indirectly from that information. It includes but is not restricted to factual information such as date of birth, ID number, or location and may include sensitive information such as health, ethnicity and opinions expressed. Once personal data has been completely anonymised so that it is impossible to re-identify the individuals concerned it ceases to be personal data. The University may use anonymised data as summary statistics or for analyses but only where it is truly anonymous.

#### DATE

___________________

[I Agree]

[I Do Not Agree]

### 3. Debrief Sheet

#### PROJECT TITLE: CAREGIVER MIND-MINDEDNESS TRAINING AS AN EARLY INTERVENTION FOR SOCIALLY ANXIOUS CHILDREN

##### INVESTIGATORS

We are a group of researchers studying children’s social anxiety. The research is supervised by Daniel Hale, Mary E. Stewart and Minu Mathews. The investigator is Hiva Javadian.

##### INTRODUCTION

Childhood social anxiety, with onset in preschool years (Hitchcock et al., 2009; Mohammadi et al., 2020), has been linked to attachment style and deficits in theory of mind (ToM) (Cristina Colonnesi et al., 2017; Ronchi et al., 2020). Caregiver-child interactions and parental mentalisation like mind-mindedness influence socio-emotional development and attachment security (Meins, 1997; Meins et al., 2003), with higher mind-mindedness promoting positive outcomes (Bernier et al., 2023; Fishburn et al., 2022; Miller et al., 2019). Culture shapes parenting and attachment styles (Kamza, 2019; Strand et al., 2019; Zaidman-Mograbi et al., 2020). COVID-19 increased child anxiety (Moore et al., 2020; Racine et al., 2021), underscoring the need for youth mental health resources. Insecure attachment and ToM deficits can contribute to social anxiety (Bowlby, 1969), while low caregiver attunement through poor mind-mindedness may exacerbate it (Meins, Centifanti, et al., 2013). Examining links between low caregiver mind-mindedness and childhood social anxiety (Meins, 2001) can inform early intervention to alleviate symptoms and prevent disorders, critical for child well-being (Courtney et al., 2020; Ng & Ng, 2022).

##### AIM OF STUDY

The aim of the study was to investigate the effectiveness of caregivers’ mind-mindedness training in reducing social anxiety symptoms among preschool-aged children. By assessing the impact of a targeted intervention on caregiver behavior and its effects on child social anxiety, this research aimed to contribute to the development of early interventions for enhancing child well-being and promoting mental health outcomes. Moreover, this study explored the potential role of attachment and theory of mind as underlying mechanisms for the observed changes in social anxiety symptoms. This study focused on how cultural differences between Iran and the UK might have moderated this relationship.

##### PROCEDURE

The study aimed to recruit 100 primary caregivers of socially anxious preschool-aged children, aged 4-7 years, from the UK and Iran. Recruitment methods included distributing flyers at preschools/nurseries, word-of-mouth, and referrals from teachers. Participants had a short max 30-minutes interview and then were randomly assigned to either the mind-mindedness training group (n=50) or the peer support group (n=50), using a computer-generated random sequence and allocation concealment to ensure unbiased assignment.

The mind-mindedness training group underwent a 3-day (1h) online training program, which included psychoeducation, emotion coaching, video examples, reflective discussions, and role-play exercises aimed at enhancing attuned, mentalizing caregiving behaviours. The peer support group had access to a private online platform for sharing experiences and coping strategies related to their child’s anxiety, without direct mind-mindedness training.

Data collection occurred at baseline, post-intervention, and 3-month follow-up using validated measures.

##### FOR FURTHER INFORMATION

Daniel Hale will be glad to answer your questions about this study and provide additional information on results if requested. You may contact them at.

##### PRIVACY AND CONFIDENTIALITY

All data will be fully anonymized during the transcription process. Any identifying details, such as names, addresses, and places of work, will be removed or destroyed to ensure confidentiality.

Heriot-Watt University is the data controller for the personal data collected in this project. We are using your personal data for this project only with your consent. You can withdraw your consent at any time until the data is fully anonymised by contacting the researcher, supervisor or the data protection team.

We will keep your personal data securely and ensure that no one will link the research data you provide to any identifying information you may supply. Once we have analysed the information you provide, we will completely anonymise your personal data so that it will not be possible to identify you from any information in the remaining dataset. After the project ends, we may use the anonymous dataset for research outputs such as articles and conference presentations.

If you would like to know more about what Heriot-Watt University does with your personal data and your rights under privacy law, please visit our data protection web pages at https://www.hw.ac.uk/uk/services/information-governance/protect/privacy-and-your-data-rights.htm or contact our Data Protection Officer by email at.

## 4. Audio Recording consent form

### FOR AUDIO RECORDING

Heriot-Watt University is the data controller for the personal data that you have consented to be collected in this project. We will keep your personal data securely and ensure that no one will link the research data you provide to any identifying information you may supply.

We are using your personal data for this project only with your consent. You can withdraw your consent at any time until the data is fully anonymised contacting the researcher, supervisor or the data protection team.

Once this recording has been transcribed or analysed, the recording will be erased and your data will be anonymised.

As a participant in this study, I agree to be audio-recorded for the purpose of collecting data for this study as well as a means of verifying results from other data collected. I am aware that I may withdraw this consent at any time without penalty, at which point, the audio recording will be erased. I understand that once this recording has been transcribed or analysed, the recording will be erased and my data will be anonymised.

## PARTICIPANT

### DATE

___________________________

[I Agree]

[I Do Not Agree]

## 5. Flyer

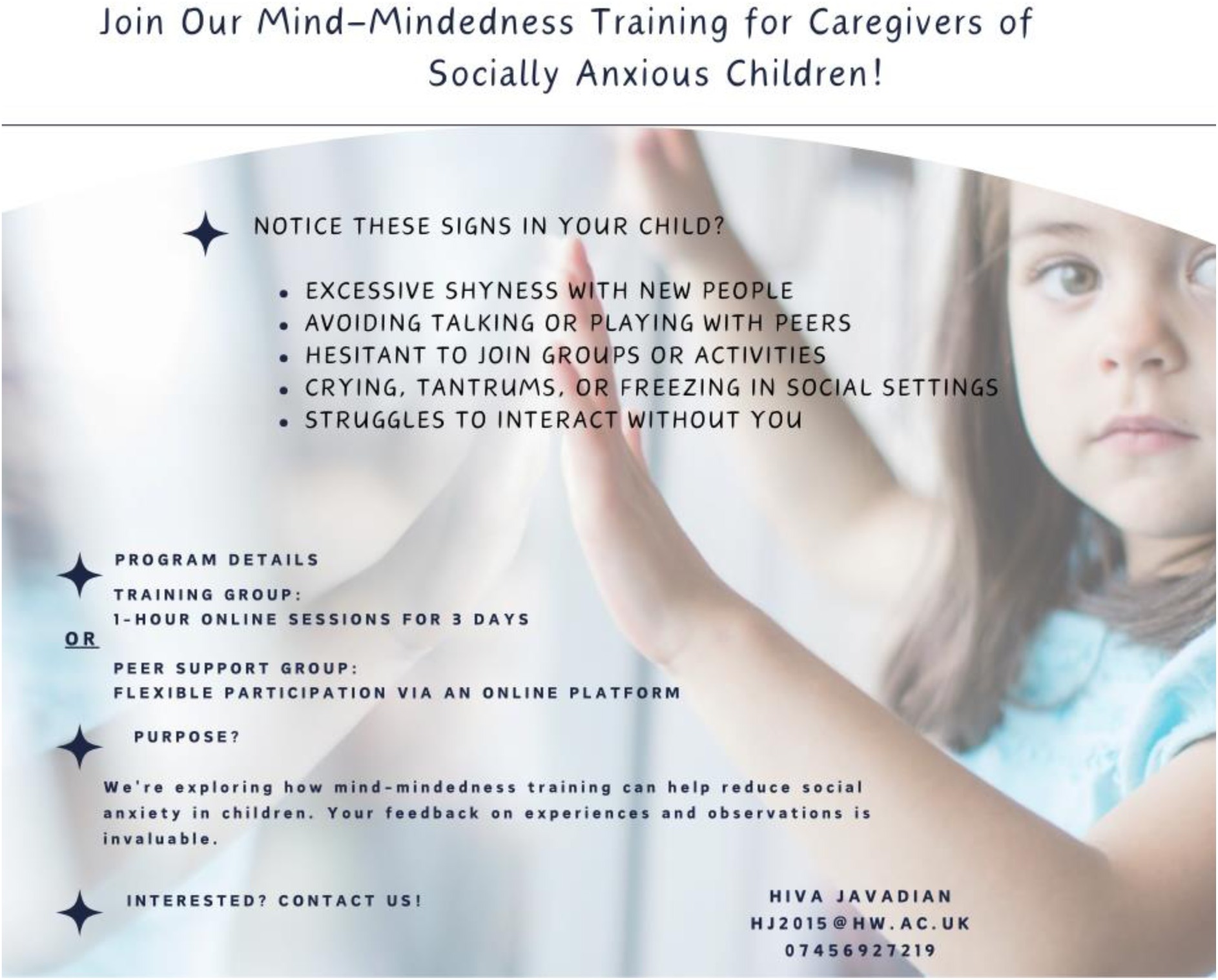

## 6. Data Collection Instruments

### 6.1 Mindmindedness Coding Manual 2.2

4. Representational Measures of Mind-Mindedness In Preschool and Older Children

In caregivers of children of preschool age and above, we have assessed MM using a brief interview (Meins, Fernyhough, Russell, & Clark-Carter, 1998). Caregivers are first informed that there are no right or wrong answers to the questions in the interview and that they should feel free to talk about the first things that come into their heads. The caregiver is simply given an open-ended invitation to describe the child: Can you describe [child’s name] for me? If caregivers seek guidance on how to answer the question, the researcher should repeat that no specific type of description is required, and that the caregiver should talk about whatever comes into his/her head. When the caregiver has finished replying, s/he is asked Can you say anything else about him/her? [If the caregiver has already given an extensive answer in reply to the first question, this prompt can be omitted.] We usually include two further follow-up questions in the MM interview (What’s the best thing about [child’s name]? and What do you try to teach [child’s name]?), but the answers to these questions are not analysed as part of the MM assessment. If the MM interview is the only measure that the caregiver will be completing in the testing session, it is useful first to put the caregiver at ease by asking general questions (e.g., whether the target child has any siblings, whether they attend preschool, their precise age, etc.) before asking the caregiver to describe the child.

Caregivers’ answers to the describe your child question are transcribed verbatim, and each attribute mentioned that refers to the child is classified into one of the four exhaustive and exclusive categories described below (Meins et al., 1998, 2003). Implicit descriptions are coded; for example, if the caregiver said ‘he wears us out’ without explicitly mentioning the relevant attribute (e.g., high activity level).

Note that, unlike in the observation-based MM coding scheme, precise repetitions of specific attributes mentioned during the interview are not coded separately, so each attribute can only be coded once. For example, if a caregiver described the child as happy twice in the interview, this would only be coded as one attribute, but if the caregiver described the child as happy and then as content, this would be coded as two attributes. The rationale for treating repetitions differently in the observation and interview MM schemes is that caregivers’ interview-based descriptions of their children are purely representational, so repeating the same mentalistic attribute does not entail a more diverse representation of the child as an individual with a mind. In contrast, mind-related comments in the observation-based scheme are in response to the infant’s behaviour, so repetitions of such comments are meaningful because they index whether the caregiver is reading the infant’s internal states appropriately or in a non-attuned manner over time.

As for the observation measure, the lead researcher should decide how to section the descriptions into individual attributes. The coders should received the descriptions in sectioned format in order to avoid confusion over what should be classified as an attribute.

### 6.2 Spence Children’s Anxiety Scale

|  |  |  |  |  |  |
| --- | --- | --- | --- | --- | --- |
| 1. | My child worries about things..... | Never | Sometimes | Often | Always |
| 2. | My child is scared of the dark..... | Never | Sometimes | Often | Always |
| 3. | When my child has a problem, s(he) complains of<br>having a funny feeling in his / her stomach ..... | Never | Sometimes | Often | Always |
| 4. | My child complains of feeling afraid..... | Never | Sometimes | Often | Always |
| 5. | My child would feel afraid of being on his/her own at home..... | Never | Sometimes | Often | Always |
| 6. | My child is scared when s(he) has to take a test..... | Never | Sometimes | Often | Always |
| 7. | My child is afraid when (s)he has to use public toilets or bathrooms..... | Never | Sometimes | Often | Always |
| 8. | My child worries about being away from us / me..... | Never | Sometimes | Often | Always |
| 9. | My child feels afraid that (s)he will make a fool of him/herself<br>in front of people..... | Never | Sometimes | Often | Always |
| 10. | My child worries that (s)he will do badly at school..... | Never | Sometimes | Often | Always |
| 11. | My child worries that something awful will happen to<br>someone in our family..... | Never | Sometimes | Often | Always |
| 12. | My child complains of suddenly feeling as if (s)he can't breathe<br>when there is no reason for this..... | Never | Sometimes | Often | Always |
| 13. | My child has to keep checking that (s)he has done things right<br>(like the switch is off, or the door is locked)..... | Never | Sometimes | Often | Always |
| 14. | My child is scared if (s)he has to sleep on his/her own..... | Never | Sometimes | Often | Always |
| 15. | My child has trouble going to school in the mornings because<br>(s)he feels nervous or afraid..... | Never | Sometimes | Often | Always |
| 16. | My child is scared of dogs ..... | Never | Sometimes | Often | Always |
| 17. | My child can't seem to get bad or silly thoughts out of his / her head..... | Never | Sometimes | Often | Always |
| 18. | When my child has a problem, s(he) complains of<br>his/her heart beating really fast..... | Never | Sometimes | Often | Always |
| 19. | My child suddenly starts to tremble or shake when there is no reason for this..... | Never | Sometimes | Often | Always |
| 20. | My child worries that something bad will happen to him/her..... | Never | Sometimes | Often | Always |
| 21. | My child is scared of going to the doctor or dentist ..... | Never | Sometimes | Often | Always |
| 22. | When my child has a problem, (s)he feels shaky..... | Never | Sometimes | Often | Always |
| 23. | My child is scared of heights (eg. being at the top of a cliff)..... | Never | Sometimes | Often | Always |
| 24. | My child has to think special thoughts (like numbers or words) to stop bad things from happening..... | Never | Sometimes | Often | Always |
| 25. | My child feels scared if (s)he has to travel in the car, or on a bus or train ..... | Never | Sometimes | Often | Always |
| 26. | My child worries what other people think of him/her..... | Never | Sometimes | Often | Always |
| 27. | My child is afraid of being in crowded places (like shopping centres, the movies, buses, busy playgrounds)..... | Never | Sometimes | Often | Always |
| 28. | All of a sudden my child feels really scared for no reason at all..... | Never | Sometimes | Often | Always |
| 29. | My child is scared of insects or spiders..... | Never | Sometimes | Often | Always |
| 30. | My child complains of suddenly becoming dizzy or faint when there is no reason for this..... | Never | Sometimes | Often | Always |
| 31. | My child feels afraid when (s)he has to talk in front of the class..... | Never | Sometimes | Often | Always |
| 32. | My child's complains of his / her heart suddenly starting to beat too quickly for no reason ..... | Never | Sometimes | Often | Always |
| 33. | My child worries that (s)he will suddenly get a scared feeling when there is nothing to be afraid of..... | Never | Sometimes | Often | Always |
| 34. | My child is afraid of being in small closed places, like tunnels or small rooms..... | Never | Sometimes | Often | Always |
| 35. | My child has to do some things over and over again (like washing his / her hands, cleaning or putting things in a certain order)..... | Never | Sometimes | Often | Always |
| 36. | My child gets bothered by bad or silly thoughts or pictures in his/her head ..... | Never | Sometimes | Often | Always |
| 37. | My child has to do certain things in just the right way to stop bad things from happening ..... | Never | Sometimes | Often | Always |
| 38. | My child would feel scared if (s)he had to stay away from home overnight..... | Never | Sometimes | Often | Always |
| 39. | Is there anything else that your child is really afraid of? ..... | YES | NO |  |  |
|  | Please write down what it is, and fill out how often (s)he is afraid of this thing: ..... | Never | Sometimes | Often | Always |
|  | ..... | Never | Sometimes | Often | Always |
|  | ..... | Never | Sometimes | Often | Always |

### 6.3 Attachment Insecurity Screening Inventory

The Children’s Social Understanding Scale (CSUS) Short Form Scoring Instructions

The Children’s Social Understanding Scale (CSUS) score is computed as follows:

Sum all item responses and divide the total by number of items receiving a score.

* If the parent skipped an item, that item receives no score and is coded as missing.
* If the parent chose “Don’t Know” as a response to an item, it is coded as missing.
* Items #4 and #10 are reverse items and must be scored in the following way:

4 becomes 1

3 becomes 2

2 becomes 3

1 becomes 4

Please cite this measure as:

Tahiroglu, D., Moses, L. J., Carlson, S. M., Mahy, C. E. V., Olofson, E. L., & Sabbagh, M. A.

(2014). The Children’s Social Understanding Scale: Construction and validation of a parent-report theory-of-mind measure. Developmental Psychology, 50, 2485-2497.Attachment Insecurity Screening Inventory (AISI) 2–5 years: 20-item Parental Report (1 = never; 2 = sometimes; 3 = regularly; 4 = often; 5 = very often; 6 = always)

1. Does your child try to force you to do what he/she wants?
2. Is your child excessively docile and obedient?
3. Does your child respond well and remain relaxed when you touch him/her (R)?
4. Does your child stay in control when playing with you?
5. Does your child enjoy being cuddled by you (R)?
6. Does your child cling to you?
7. Does your child argue with you if things do not turn out the way he/she expects?
8. Does your child let you comfort him/her if he/she is in pain, frightened or upset (R)?
9. Does your child ask for help with problems (R)?
10. Is your child over-concerned when you are upset or unwell?
11. Is it easy for your child to resume contact with you after you have been separated (R)?
12. Is your child excessively determined to decide everything for him/herself?
13. Is your child excessively emotional when you leave him/her for a short period of time?
14. Is your child able to enjoy contact with you (R)?
15. Does your child want to be put down and then immediately picked up again?
16. Does your child keep a close eye on you while you do things in and around the house?
17. Does your child hug or cuddle you spontaneously (R)?
18. Does your child become angry with you quickly?
19. Is your child happy and playful in your presence (R)?
20. Does your child need you to reassure him/her that he/she is doing something right?

Note. Avoidance subscale items: 3, 5, 8, 9, 11, 14, 17, 19, Ambivalence/resistance subscale

items: 2, 6, 10, 13, 15, 16, 20, Disorganization subscale items: 1, 4, 7, 12, 18; (R) = reversely coded.

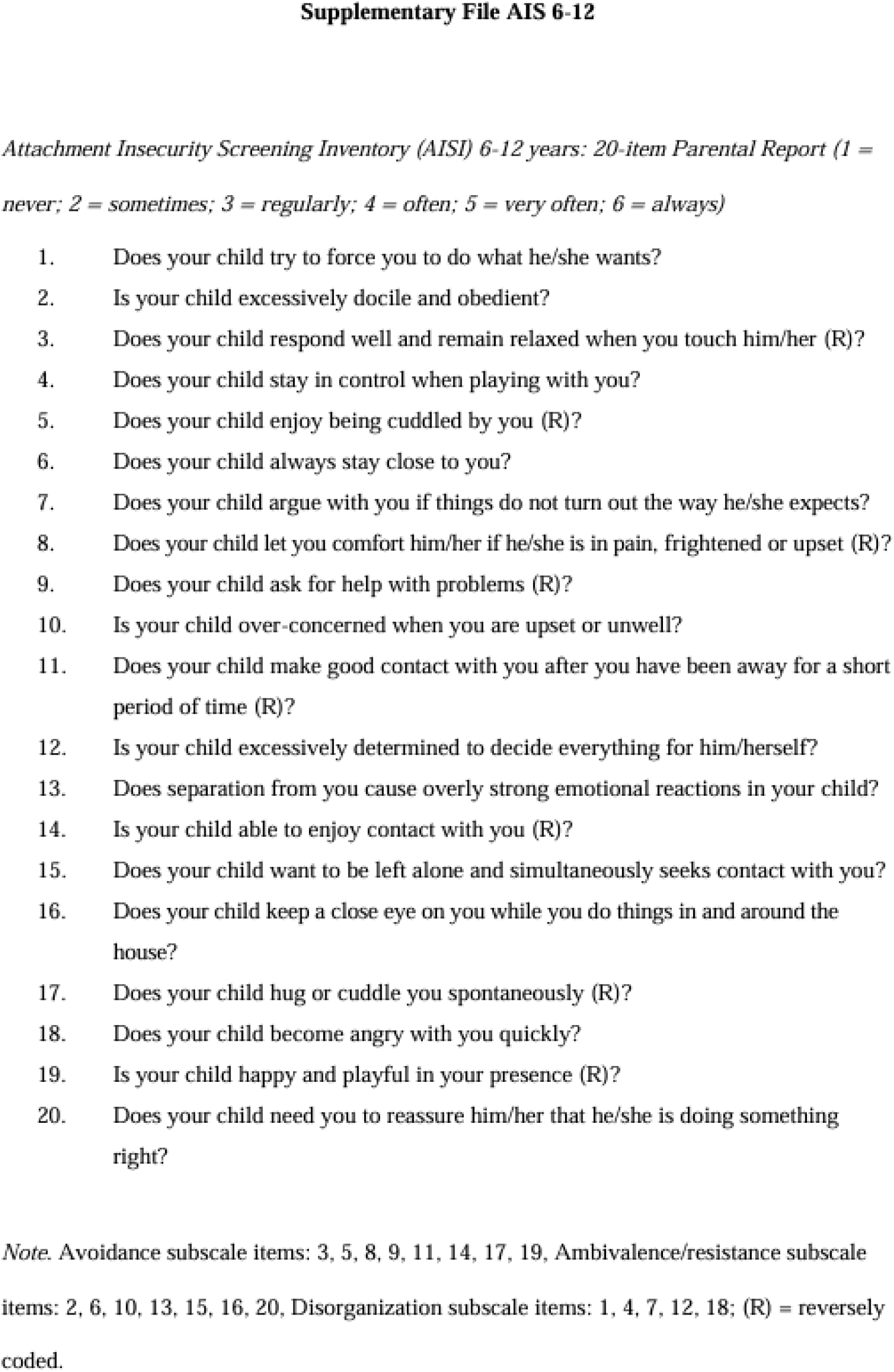

### 6.4 Theory of Mind Task Battery

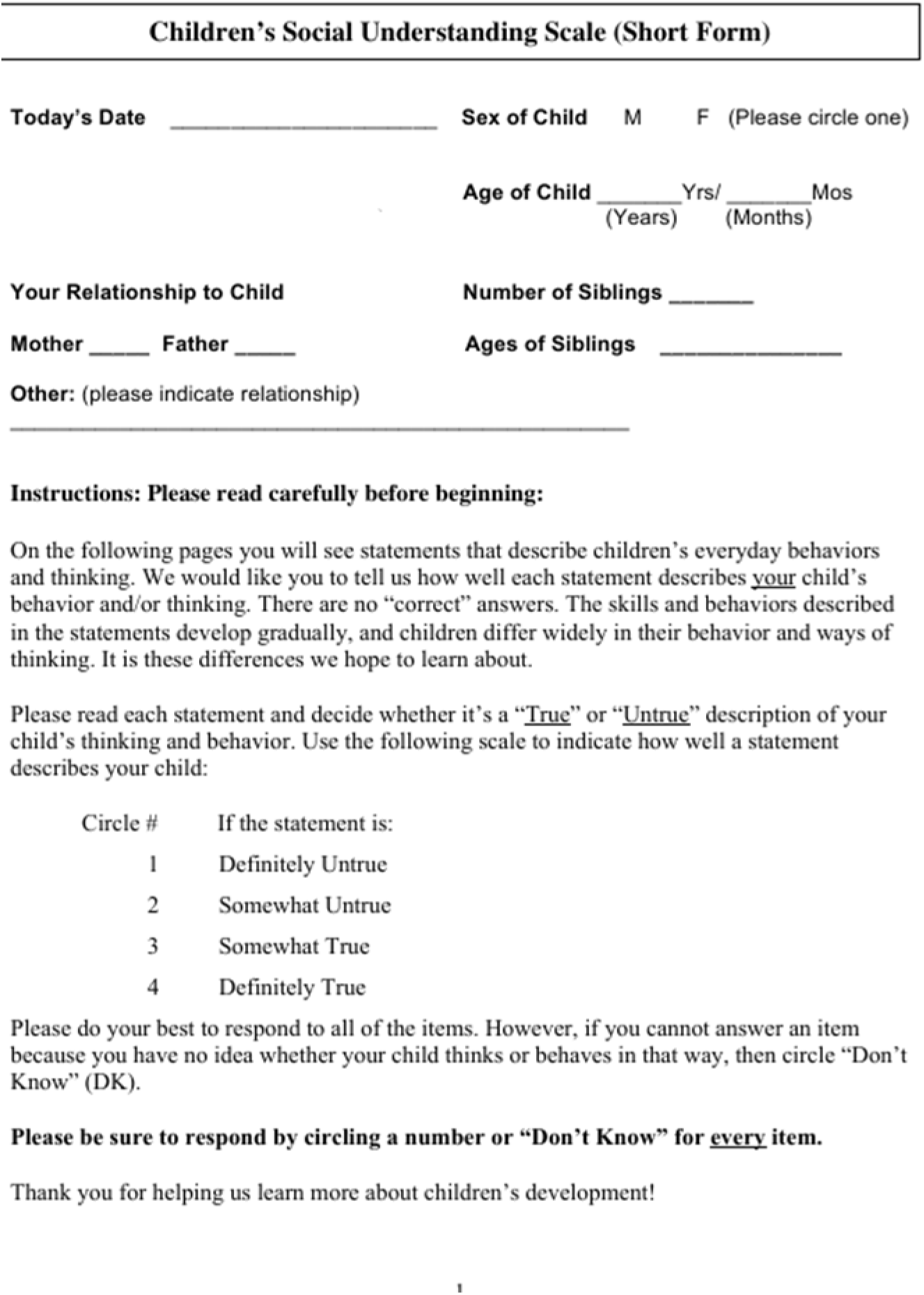
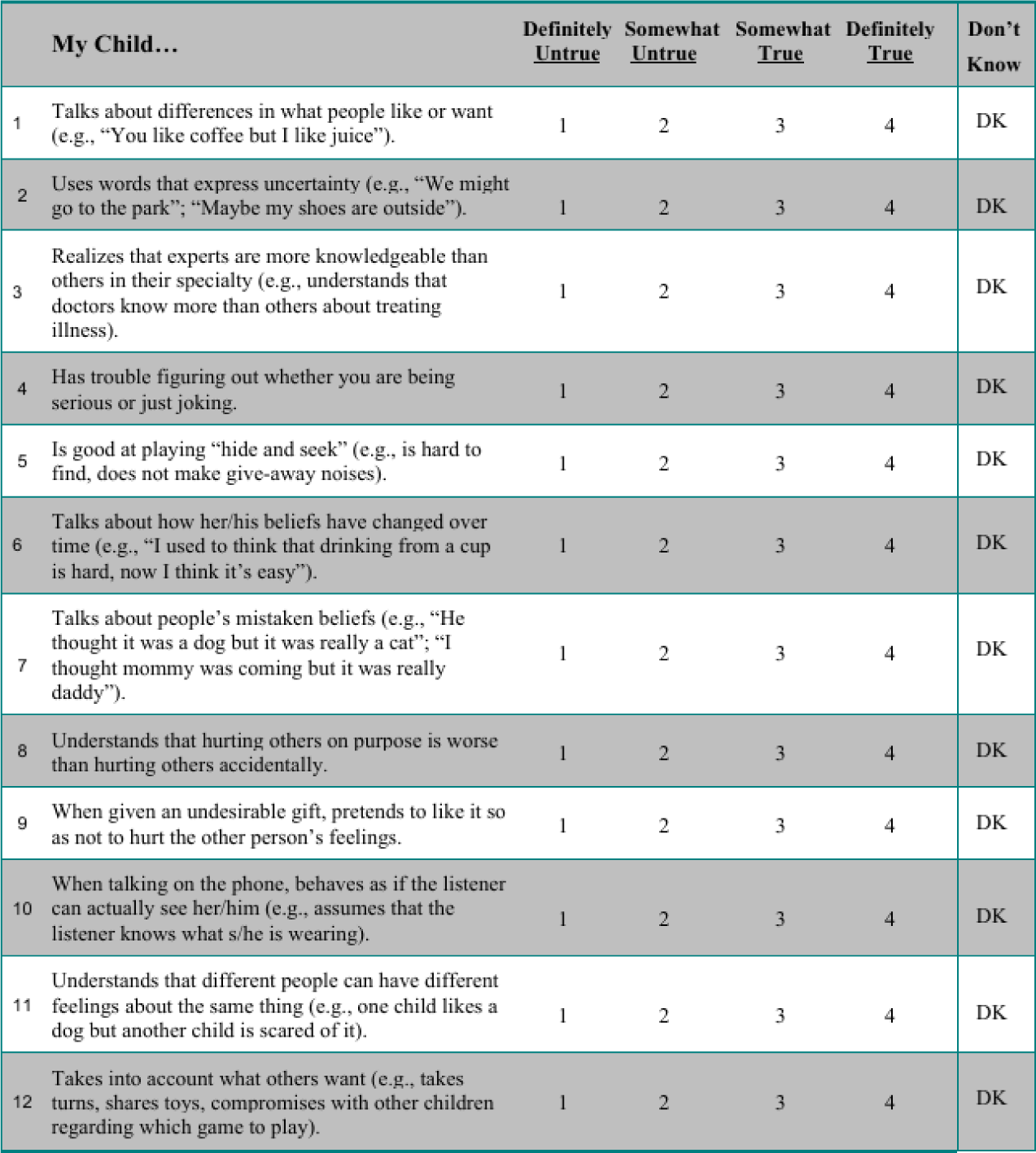
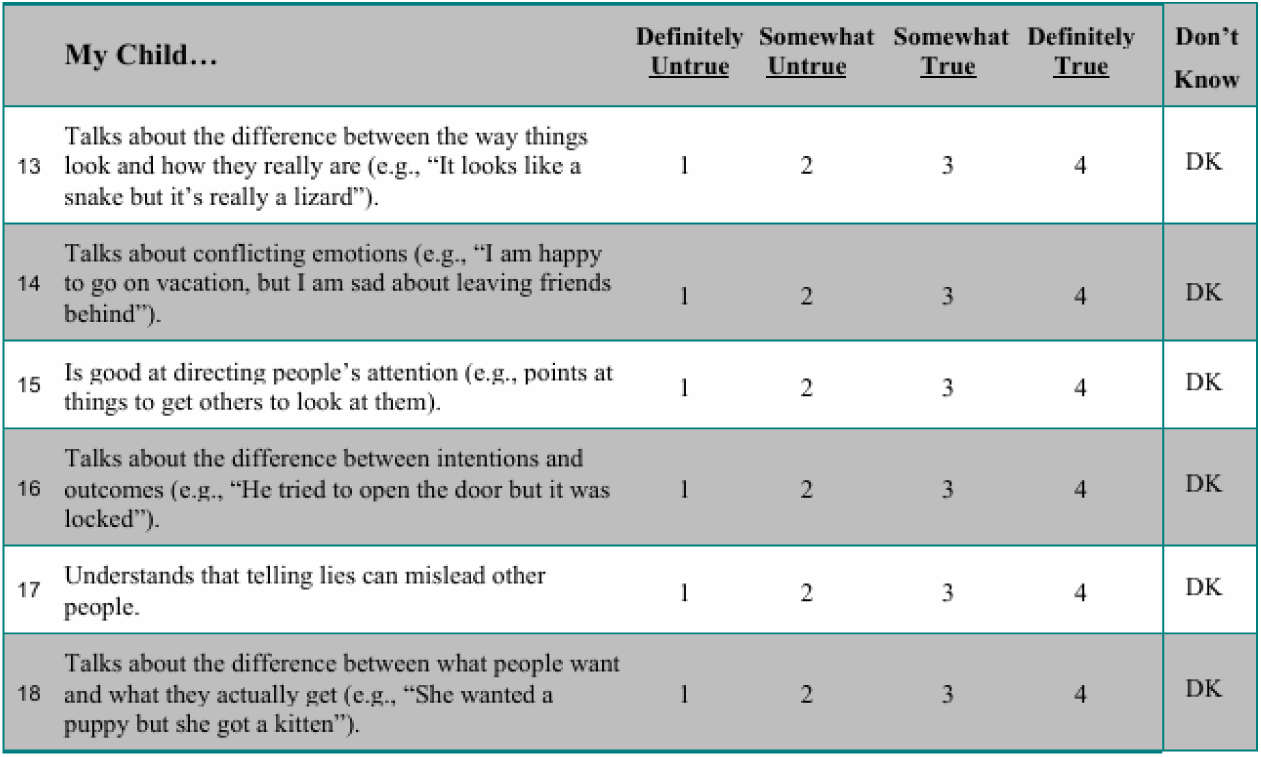

| Children's Social Understanding Scale (Short Form) |  |
| --- | --- |
| Today's Date _____ | Sex of Child M F (Please circle one) |
| Age of Child _____ Yrs/ _____ Mos<br>(Years) (Months) |  |
| Your Relationship to Child | Number of Siblings _____ |
| Mother _____ Father _____ | Ages of Siblings _____ |
| Other: (please indicate relationship)<br>_____ |  |
| 1 | Definitely Untrue |
| 2 | Somewhat Untrue |
| 3 | Somewhat True |
| 4 | Definitely True |

|  | My Child... | Definitely<br>Untrue | Somewhat<br>Untrue | Somewhat<br>True | Definitely<br>True | Don't<br>Know |
| --- | --- | --- | --- | --- | --- | --- |
| 1 | Talks about differences in what people like or want (e.g., "You like coffee but I like juice"). | 1 | 2 | 3 | 4 | DK |
| 2 | Uses words that express uncertainty (e.g., "We might go to the park"; "Maybe my shoes are outside"). | 1 | 2 | 3 | 4 | DK |
| 3 | Realizes that experts are more knowledgeable than others in their specialty (e.g., understands that doctors know more than others about treating illness). | 1 | 2 | 3 | 4 | DK |
| 4 | Has trouble figuring out whether you are being serious or just joking. | 1 | 2 | 3 | 4 | DK |
| 5 | Is good at playing "hide and seek" (e.g., is hard to find, does not make give-away noises). | 1 | 2 | 3 | 4 | DK |
| 6 | Talks about how her/his beliefs have changed over time (e.g., "I used to think that drinking from a cup is hard, now I think it's easy"). | 1 | 2 | 3 | 4 | DK |
| 7 | Talks about people's mistaken beliefs (e.g., "He thought it was a dog but it was really a cat"; "I thought mommy was coming but it was really daddy"). | 1 | 2 | 3 | 4 | DK |
| 8 | Understands that hurting others on purpose is worse than hurting others accidentally. | 1 | 2 | 3 | 4 | DK |
| 9 | When given an undesirable gift, pretends to like it so as not to hurt the other person's feelings. | 1 | 2 | 3 | 4 | DK |
| 10 | When talking on the phone, behaves as if the listener can actually see her/him (e.g., assumes that the listener knows what s/he is wearing). | 1 | 2 | 3 | 4 | DK |
| 11 | Understands that different people can have different feelings about the same thing (e.g., one child likes a dog but another child is scared of it). | 1 | 2 | 3 | 4 | DK |
| 12 | Takes into account what others want (e.g., takes turns, shares toys, compromises with other children regarding which game to play). | 1 | 2 | 3 | 4 | DK |
| 13 | Talks about the difference between the way things look and how they really are (e.g., "It looks like a snake but it's really a lizard"). | 1 | 2 | 3 | 4 | DK |
| 14 | Talks about conflicting emotions (e.g., "I am happy to go on vacation, but I am sad about leaving friends behind"). | 1 | 2 | 3 | 4 | DK |
| 15 | Is good at directing people's attention (e.g., points at things to get others to look at them). | 1 | 2 | 3 | 4 | DK |
| 16 | Talks about the difference between intentions and outcomes (e.g., "He tried to open the door but it was locked"). | 1 | 2 | 3 | 4 | DK |
| 17 | Understands that telling lies can mislead other people. | 1 | 2 | 3 | 4 | DK |
| 18 | Talks about the difference between what people want and what they actually get (e.g., "She wanted a puppy but she got a kitten"). | 1 | 2 | 3 | 4 | DK |

## 7. Ethics Approval Letter

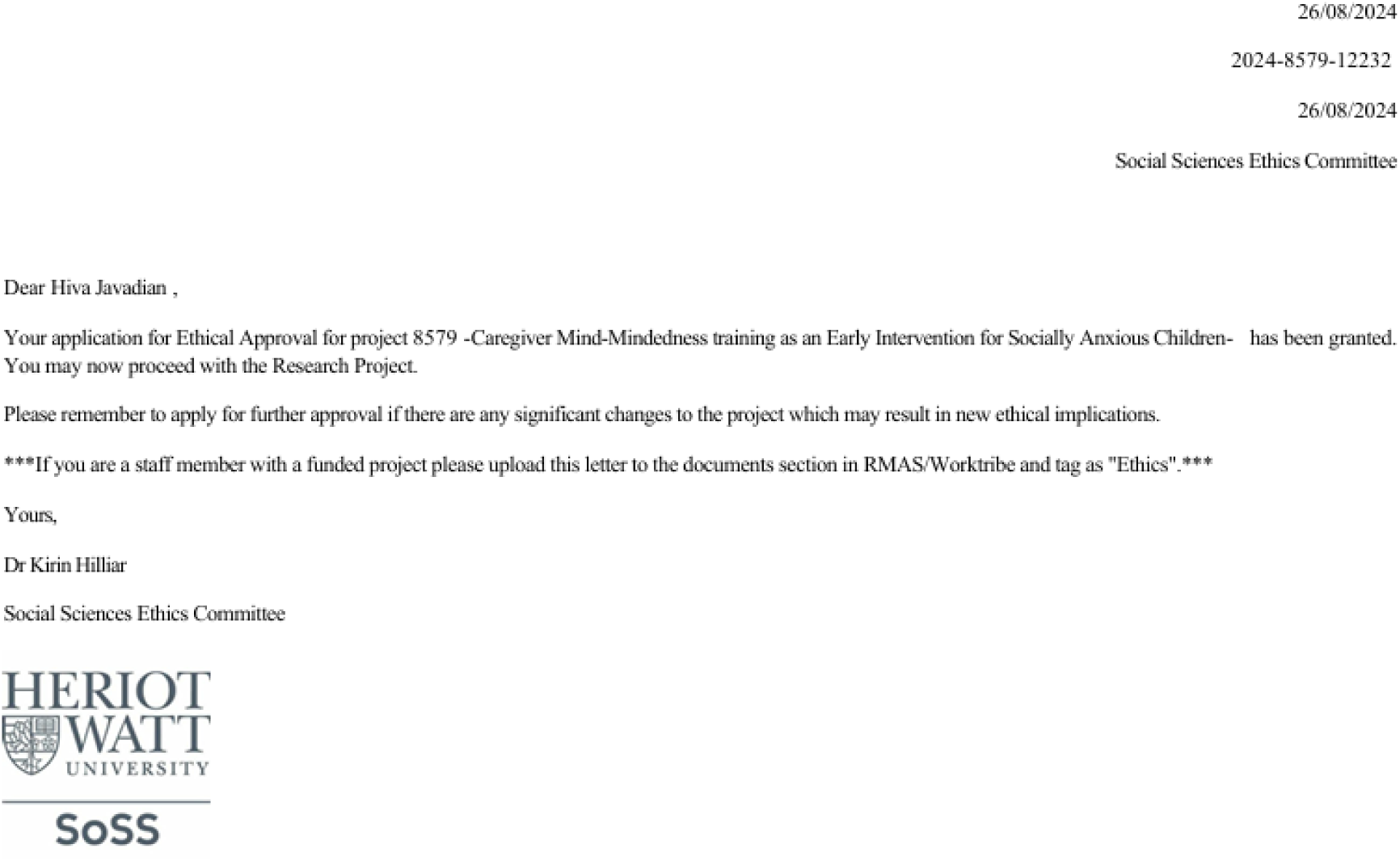

## Notes

### Competing Interest Statement

The authors have declared no competing interest.

### Clinical Trial

NCT Number NCT06657014 Iranian Registry of Clinical Trials (IRCT) ID: 80088

### Funding Statement

The author(s) received no specific funding for this work.

### Author Declarations

Name of IRB/Ethics Committee: Social Sciences Ethics Committee Approval Number: 2024-8579-12232 Approval Statement: This study was approved by the Social Sciences Ethics Committee (Approval No: 2024-8579-12232) on 26/08/2024.

